# Leg ulcers are associated with an increased risk of Sickle eye disease in Lagos

**DOI:** 10.1101/2022.04.22.22274153

**Authors:** Olubanke Theodora Ilo, Olufemi Emmanuel Babalola, Kuburat Oliyide, Michael Olufemi Kehinde, Folasade Akinsola

## Abstract

**Key Point:** **Patients with sickle cell Leg ulcers are 5.9 times more likely to develop sickle retinopathy**.

Leg ulcers are a common sign in Sickle Cell Disease. But to what extent are they indicative of Sickle cell eye disease in these patients? This short communication assesses to what extent leg ulcers predict the presence of sickle eye disease in Nigerian patients.

This was a clinic based, comparative non-interventional study, conducted at both the Hematology Clinic and Eye clinic in Lagos University Teaching Hospital (LUTH). One hundred consecutive cases of Sickle Cell Disease (SCD) patients (HbSS and HbSC) attending the Haematology Clinic were compared with 100 age and sex matched non-SCD HbAA controls. The presence of leg ulcers was sought in both groups, and all had assessment for ocular anterior and posterior signs of SCD. Cases and controls were comparable in age and sex. Amongst 100 Cases, 85 were HbSS and 15 HbSC. Leg ulcers were present in 18 patients. Eighty-two cases had either anterior or posterior SCD related ocular manifestations. Patients with leg ulcer were found to be at higher risk of developing Conjunctival Sickle Sign, Retinal Venous Tortuosity and Proliferative Sickle Retinopathy with an increased likelihood of 22.9, 7.1 and 5.9 times respectively. We believe this is the first study to quantify this association.

**Conclusions:** This study suggests that sickle cell eye disease is more likely to develop in sickle cell disease patients with leg ulcers. Internists managing SCD patients are advised to refer patients with leg ulcers for early ocular assessment in order to prevent complications.

## INTRODUCTION

Sickle cell disease refers to a group of genetic disorders characterized by the presence of sickled red blood cells, anaemia, as well as acute and chronic tissue injury secondary to blockage of blood flow by abnormally shaped red blood cells^1^. The rigid sickled erythrocytes cause disruption of blood flow leading to vascular occlusions, hemorrhage, infarctions and ischaemic necrosis of tissues and organs in the body including the eyes. The disease is found almost exclusively in blacks^2^ with no gender predilection.

The ocular manifestations of sickle cell disease result from vascular occlusion, which may occur in the conjunctiva, iris, vitreous, retina, and choroid^3^. The occurrence of these ophthalmic manifestations have been well documented in literature^3-5^.

However less documented and elucidated are attendant risk factors that could predispose one developing sickle cell eye disease in adult patients living with sickle cell disease. As treatment modalities improve, individuals with sickle cell disease live longer thus increasing the incidence of sickle cell related eye disease^6,7^. With Nigeria having the largest population of people with sickle cell disorder in the world^8^, more of the population is at risk of the blinding complications and other ocular problems. There is therefore a need for more awareness on the ocular morbidities associated with sickle cell disease, and the elucidation of factors that could point to potential blinding disease.

Leg ulceration occur in SCD, especially in the monozygotic HBSS disease^9,10,11^ The mechanism may include mechanical obstruction of blood flow by logjammed sickled cells, and decreased nitric acid bioavailability ^12^

Leg ulcers are most commonly found in areas of the body with low subcutaneous fat such as the medial and lateral malleolus^13^

Akinyanju and Akinsete ^10^ reported a leg ulceration prevalence of 1.4% in SCD patients in Lagos and indicated that it tended to be commoner in males than females.

We recently carried out a study in Lagos in which ocular and physical changes in sickle cell disease patients were simultaneously observed. In this short communication, we wish to report on the relationship between leg ulcers and ocular disease, and determine to what extent if any leg ulcers are a risk factor for ocular disease.

## METHODS

Ethical approval was obtained from the Lagos University Teaching Hospital (LUTH) Ethics and Research Committee (ADM/DCST/HREC/372), adhering to the tenets of the Declaration of Helsinki^14^.

The study was conducted at both the Haematology Clinic and Guinness Eye Centre (GEC) of the Lagos University Teaching Hospital (LUTH) Idi-Araba Lagos, which is the largest referral center in the region, between February 29 and April 11 2012. The cases were made up of sickle cell disease patients (genotypes HbSS, HbSC) aged 16 years and over, while the controls were age and sex matched patients with genotype HbAA drawn from consenting accompanying relatives.

Patients with systemic co-morbidities such as diabetes mellitus, hypertension, HIV/AIDS, lymphomas, and bleeding disorders like hemophilia were excluded from the study.

All the study subjects were counseled individually on the nature of the study and informed written consent obtained before proceeding with the study. Detailed Systemic and ocular examination were conducted by a consultant hematologist and then ophthalmologist. All participants were examined with a slit lamp biomicroscope at 12x magnification for anterior segment manifestations. Dilated fundoscopy was done followed by binocular indirect ophthalmoscopy to examine the fundus of all participants. Grading of sickle cell retinopathy was according to Goldberg’s classification^15^. Selected cases had pictures taken of their anterior segment manifestations and fundi. Pictures were taken of some cases of leg ulcers in the same series. (Figures 1-4)

**Figure 1:**
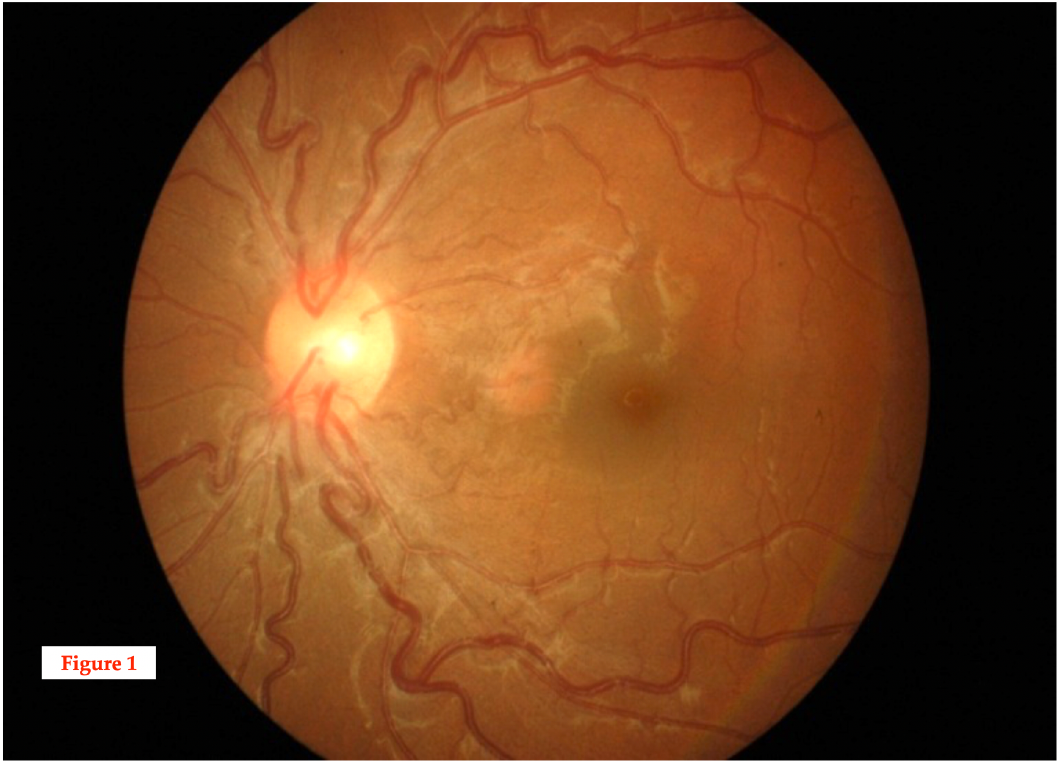
FUNDUS PHOTO SHOWING RETINAL VASCULAR TORTOUSITY IN A SUBJECT WITH SCD

**Figure 2:**
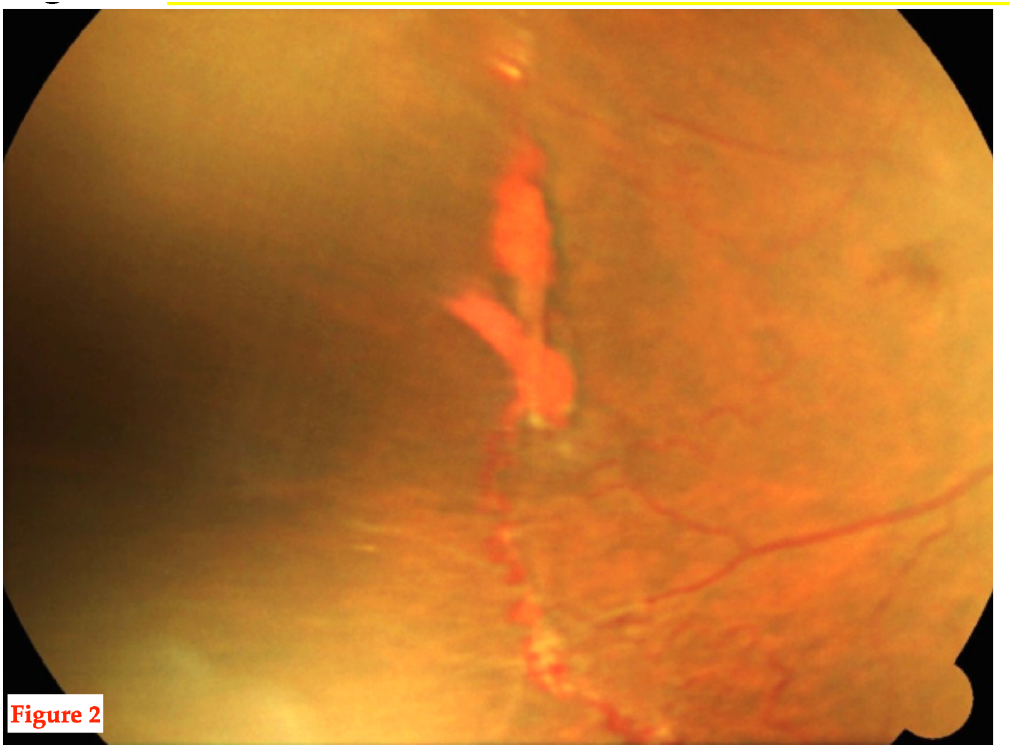
FUNDUS PHOTO SHOWING SEA FAN NEOVASCULARIZATION AND HEMORRHAGE”

**Figure 3:**
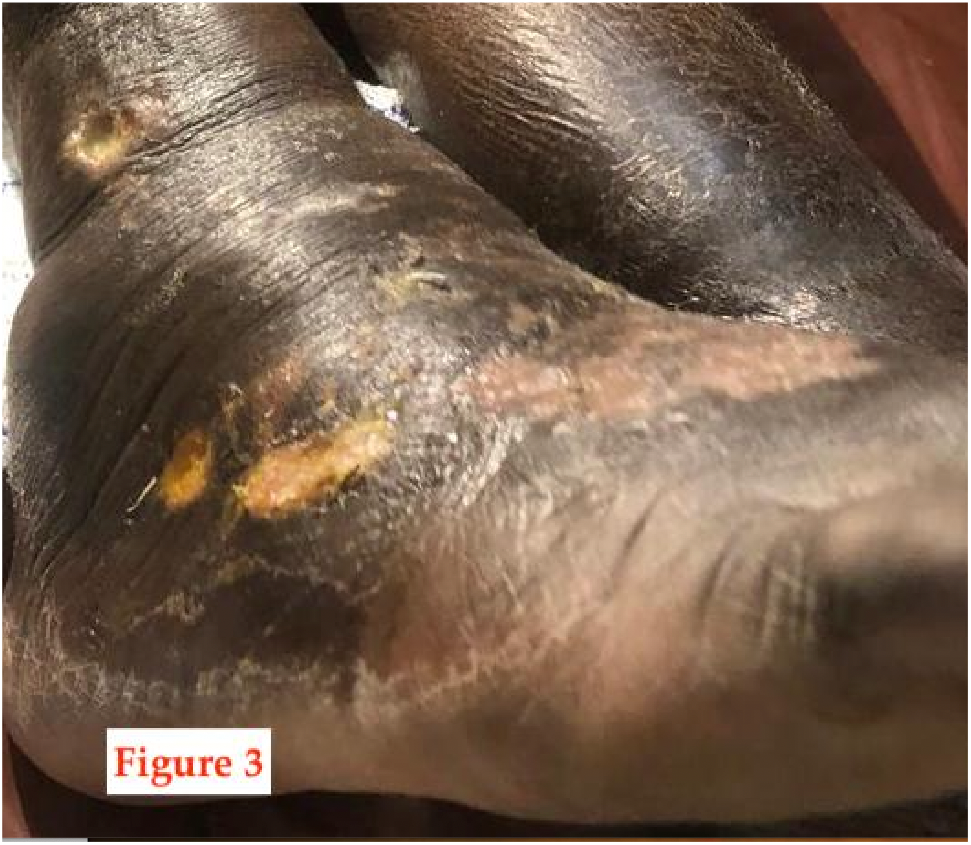
LEG ULCER INVOLVING ANKLE AND DORSUM IN A SUBJECT WITH SCD”

**Figure 4:**
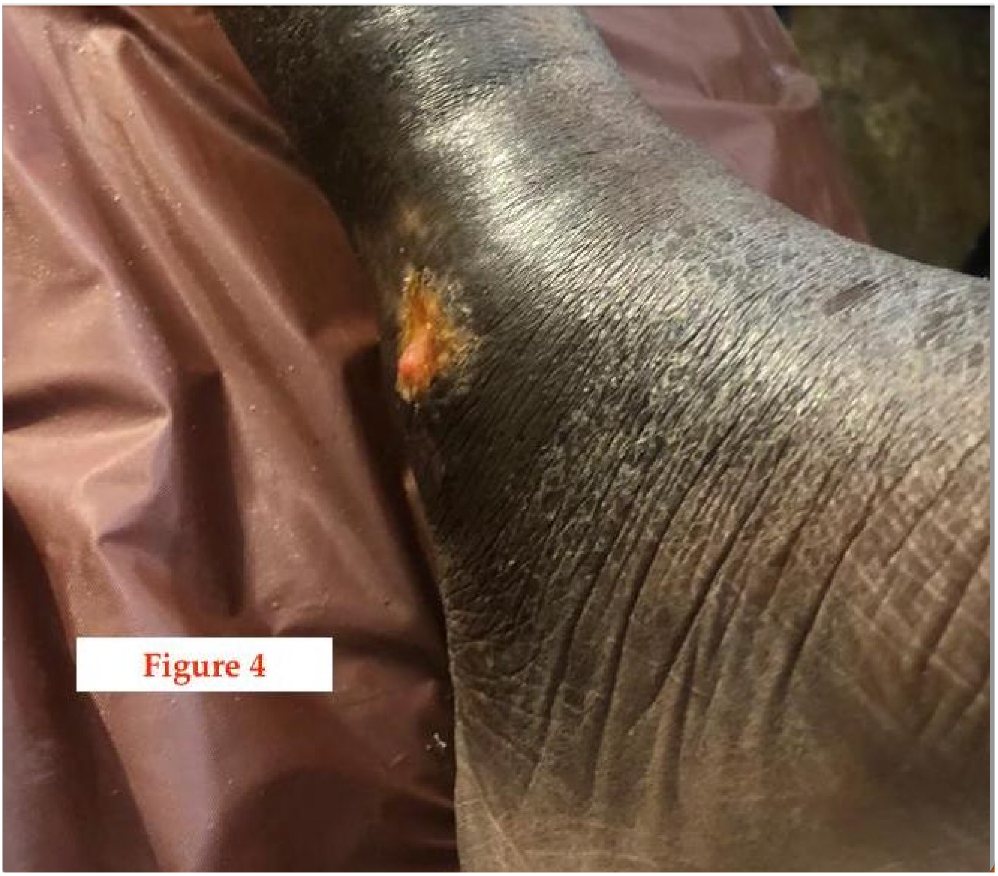
MEDIAL MALLEOLUS LEG ULCER IN A SUBJECT WITH SCD

Findings were collated and analyzed with SPSS 20. Means were compared using the student t test. Chi square and simple logistic regression were done to measure the strength of association.

## RESULTS

A total of 200 subjects participated in the study, 100 cases with sickle cell disease (85 HbSS and 15 HbSC) and 100 HbAA (controls). The two groups were statistically comparable in terms of age and sex. Age range was 16-58 years: mean age 27.55±8.53 (cases) 26.85±8.77 (controls). Females slightly outnumbered males (Male: female 1:1.4)

Distribution of visual acuity was similar in both groups (Table 1). 5 and 6 patients respectively in the case and control group had VA less than 6/18, while only 2 of the cases had VA less than 3/60 and were considered blind by World Health Organisation criteria.

**Table 1:** Visual acuity in 200 cases and controls.

| Visual acuity | Cases<br>N (%) | Controls<br>N (%) |
| --- | --- | --- |
| 6/5-6/12 | 90 (90) | 92 (92) |
| <6/12-6/18 | 3 (3) | 2 (2) |
| <6/18-3/60 | 5 (5) | 6 (6) |
| <=3/60 | 2 (2) | 0 (0) |
| Total | 100 (100) | 100 (100) |

Eighty-two of 100 cases had either anterior or posterior ocular manifestations compatible with sickle cell disease which was found in none of the controls.

Conjunctival Sickle Sign (CSS) was present in 63 cases while controls had none.

Typical Non-proliferative posterior segment lesions were found in the cases and included Retinal Vascular Tortuosity RVT (17), Salmon patch hemorrhage (1), Black sunburst (9) and Angioid Streak (1). Only 1 case of what appeared to be RVT was found in the control group. Table 2

**Table 2:** Non-Proliferative Retinopathy in cases and controls.

| Variable | Cases<br>N (%) | Controls<br>N (%) | Total |
| --- | --- | --- | --- |
| Retinal<br>vascular<br>tortuosity | 17 (17) | 1 (1) | 18 (9) |
| Salmon patch<br>hemorrhage | 1 (1) | 0 | 1 (0.5) |
| Black<br>sunburst | 9 (9) | 0 | 9 (4.5) |
| Angioid<br>streak | 1 (1) | 0 | 1 (0.5) |

Proliferative Sickle Retinopathy was present in 9 of the 100 cases, but none of the controls. However, it was more prevalent in the SC disease arm than the SS arm (25% versus 7.6%) (Table 3) **OR 3.29 (95% CI 0.73-14.95), P=0.06**. This is just short of statistical significance, probably because of low numbers, but it suggests that patients with SC diseases are more than thrice as likely to develop PSR than SS patients.

**Table 3:** Proliferative sickle cell retinopathy (PSR) by genotype SC and SS.

| **Any Retinopathy** | **SS** | **SC** | **Total** |
| --- | --- | --- | --- |
| **None** | **79** | **12** | **91** |
| **Present** | **6 (7.6%)** | **3(25%)** | **9 (9%)** |
| **Total** | **85** | **15** | **100** |

**OR 3.29 (95% CI 0.73-14.95), P=0.06.**

None of the signs of Posterior Sickle retinopathy were found in the control Arm (AA).

Leg ulcers were found in 18 individuals out of the total of 200 in the study. (Figures 1c and d). As indicated in table 4, leg ulcers were proportionately commoner in patients with Conjunctival Sickle Sign, Retinal Vascular Tortuosity, and sickle Cell Retinopathy in general. The Odds ratios were 22.98, 7.08 and 5.87 respectively (see table 4 for details), all of which were statistically significant.

**Table 4:**
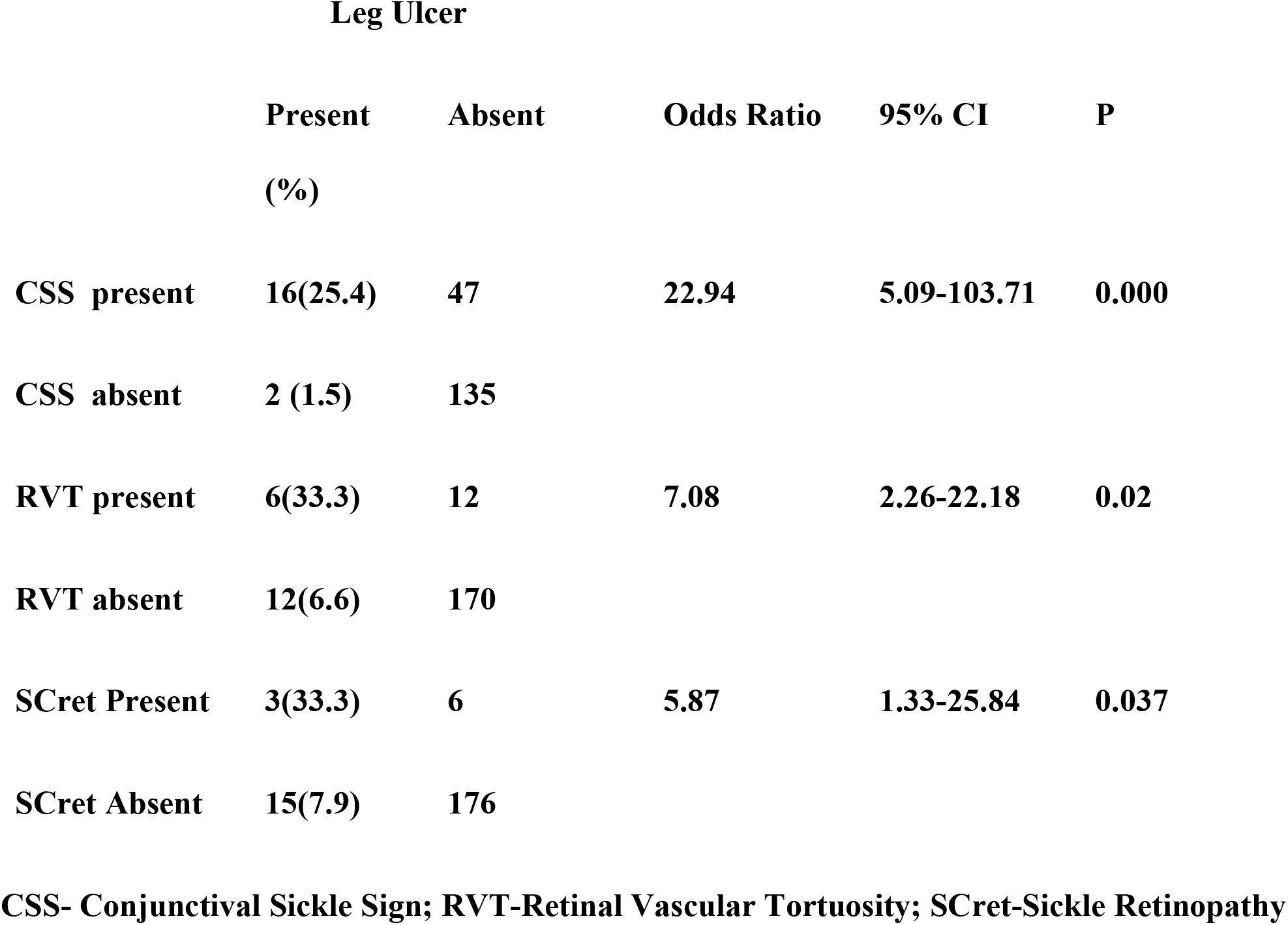
Distribution of leg ulcers and ocular pathologies.

|  | Leg Ulcer |  | Odds Ratio | 95% CI | P |
| --- | --- | --- | --- | --- | --- |
|  | Present<br>(%) | Absent |  |  |  |
| CSS present | 16(25.4) | 47 | 22.94 | 5.09-103.71 | 0.000 |
| CSS absent | 2 (1.5) | 135 |  |  |  |
| RVT present | 6(33.3) | 12 | 7.08 | 2.26-22.18 | 0.02 |
| RVT absent | 12(6.6) | 170 |  |  |  |
| SCret Present | 3(33.3) | 6 | 5.87 | 1.33-25.84 | 0.037 |
| SCret Absent | 15(7.9) | 176 |  |  |  |

**CSS- Conjunctival Sickle Sign; RVT-Retinal Vascular Tortuosity; SCret-Sickle Retinopathy**

## DISCUSSION

The link between leg ulceration and sickle cell eye disease has been drawn in this study, particularly to the effect that retinopathy is six to seven times commoner in patients with leg ulceration. This, as far as we can tell, is the first time this link will be made, even if it is logically to be expected. This is important because physicians who would generally take care of patients with leg ulcers are not often equipped or inclined to examine the back of the eye, where potentially blinding complications can occur. Early referral to ophthalmologists can lead to sight -saving intervention with the use of Pan-Retinal Laser photocoagulation or anti-Vascular Endothelial Growth Factor (VEGF) therapy, systemic therapy with hydroxycarbamide or hydroxyurea, exchange transfusion, and hyperbaric oxygenation. Surgical management such as vitrectomy and retinal detachment surgery may be indicated for recalcitrant vitreous hemorrhage. However, exoplants should be used with caution to obviate the risk of retinal ischemia^16^.

By the same token, ophthalmologists who tend to focus on eye examination alone are encouraged to examine patients more holistically, particularly for leg ulcers, so that appropriate intervention may be instituted upon referral.

There are probably overlapping pathways in the development of Sickle retinopathy and leg ulcers. Trent and Krisner ^17^ have hypothesized on various mechanisms for the development of skin ulcers in sickle cell disease. These include microvascular occlusion, secondary inflammation through release of pro-inflammatory cytokines, platelet aggregation, pro-coagulant cascade, ischemic injury and end organ damage. De Oliviero et al^18^ also hypothesized that oxidative stress plays a role in the pathogenesis of ulcers, especially in patients with glutathione S-transferase polymorphism (GSTM1 and GSTT1 null phenotypes). Other mechanisms include autonomic dysfunction of microvascular circulation^19^. and bacterial colonization involving the ulcer skin microbiome^20^. As yet poorly understood genetic factors also likely play a role, given the wide geographical variability in the incidence of the disease^21^

By the same token, the pathogenesis of Sickle cell disease ocular disorders has been assessed by Emmerson and Lutty^22^ and Walton et al^23^. The normal malleability of red blood cells RBCs allow them to pass through smaller blood vessels especially capillaries. In SCD, the RBCs take up a rigid, sickle shape in hypoxic conditions due to conversion of soluble hemoglobin to crystalline hemoglobin, an irreversible change. In addition, there is an increase in the adhesive properties of the capillary endothelium which further decreases vascular flow and causes occlusion of the vessels. The log-jamming of end vessels in the eye cause the characteristic damage seen in SCD. Where vaso-proliferative changes supervene, the changes called Proliferative Sickle Retinopathy are seen, with hypoxia and secondary hemorrhage.

The management of leg ulcers is unfortunately, not straightforward. Management generally includes the optimal hematological care of the condition, pain management, compression therapy, review of walking and footwear, and regular assessment of tissue viability^24^. Nwagu et al^25^ also advocated the use of Honey combined with transfusion with HbAA blood in the management of the condition.

The earlier these treatment modalities are instituted, the better the prognosis for skin ulcer care. One important difference of course, is that while skin ulcers are commoner in homozygous HbSS disease^26^, while retinopathy is commoner in HbSC disease^27,28^

In summary, because of the increased risk of visual complications in patients with sickle leg ulcers, it is important for attending physicians to refer patients with leg ulcers early for ophthalmological assessment. Equally, ophthalmologists should be on the lookout for leg ulcers when they are assessing SCD patients for eye disease, so that early intervention can be brought about.

## Data Availability

All data produced in the present study are available upon reasonable request to the authors

## RECOMMENDATIONS

1. It is recommended that sickle cell disease patients especially HbSC should have full medical examination including routine eye examination to optimize their eye care - for early diagnosis and prompt treatment of blinding complications.
2. Multidisciplinary approach to the management of sickle cell disease through a regular communication between the ophthalmologists and other health care givers, especially Haematologists, should be encouraged.
3. Haematologists and all doctors attending to sickle cell disease patients with leg ulcers should refer these patients for eye examination.

## AUTHORS CONTRIBUTION

Olubanke Theodora Ilo^*^ (1^st^ Author),

Olufemi Emmanuel Babalola (Co-Author),

Kuburat Oliyide (Co-Author),

Michael Olufemi Kehinde (Co-Author),

Folasade Akinsola (Co-Author).

## LIST OF ABBREVIATIONS

BIO: Binocular indirect ophthalmoscopy
CSS: Conjunctival sickle sign
CI: Confidence interval
GEC: Guinness Eye Centre
HbAA: Hemoglobin AA
HbAS: Hemoglobin AS
HbAC: Hemoglobin AC
HbSS: Hemoglobin SS
HbSC: Hemoglobin SC
LUTH: Lagos University Teaching Hospital
OR: Odd’s ratio
PSR: Proliferative sickle retinopathy
RVT: Retinal vascular tortuosity

